# Childhood socioemotional and cognitive development and Adolescents NEET (not in education, employment or training): findings from the UK Millennium Cohort Study

**DOI:** 10.1101/2025.10.29.25339038

**Authors:** Lateef Akanni, Michelle Black, Kalu Udu, Yanhua Chen, Rosalie Cattermore, Oluwaseun B. Esan, Hanna Creese, G J Melendez-Torres, Dougal Hargreaves, Nicholas Kofi Adjei, David C Taylor-Robinson

**Affiliations:** Department of Public Health, Policy and Systems, University of Liverpool, Liverpool-United Kingdom; School of Public Health, Imperial College London, London, UK; University of Exeter. Exeter, Devon, UK

**Keywords:** Cognitive ability, Socioemotional behaviour, Developmental trajectories, Education, Employment, Training, NEET, Population attributable fractions

## Abstract

**Background:** There is a growing concern about the increasing number of young people who are not in employment, education or training (NEET) globally. This study investigates the impact of concurrent cognitive and socioemotional development trajectories in childhood on NEET status in adolescence in a UK cohort.

**Method:** We analysed longitudinal data on 8,368 children from the UK Millennium Cohort Study. Exposure trajectories of cognitive and socioemotional development from age 3 to 14 years were characterised using group-based multi-trajectory models. We used Poisson regression to examine associations between developmental trajectories and NEET status at age 17, adjusting for confounders. Population-attributable fractions were estimated to quantify NEET proportions attributable to the developmental problems.

**Results:** At age 17, 3.5% of participants were NEET; of which about one-third (38%) were not economically active. Children with persistent cognitive and socioemotional development problems had a fourfold increased risk of being NEET (adjusted risk ratio [ARR] 4.0; 95% CI 2.5–6.3), and those with late socioemotional problems had threefold increased risk (3.3; 95% CI 2.2–4.9), compared to children in the no problem group. Early and resolving socioemotional and cognitive problems were not associated with being NEET. An estimated 28% (95% CI 18% to 36%) of NEET cases were attributable to cognitive and socioemotional behaviour problems in childhood.

**Conclusion:** Childhood cognitive and socioemotional development play a critical role in shaping pathways to education and employment in adolescence. Thus, policies and strategies aiming to reduce NEET should target early social and emotional skills, alongside efforts to support academic achievement.

**Strengths and limitations of this study:**

▪ The study uses longitudinal data from a contemporary and representative cohort of UK children.
▪ The study combines measures of cognitive ability and socioemotional behaviour during childhood, and evaluates the joint effects on youth NEET status
▪ A major limitation was inability to capture transitions in the NEET status as it was measured at a single time point
▪ As with most longitudinal cohort studies, missing data is inevitable and hence a challenge for analysis.

## Introduction

Education, training and employment are key gateways to human capital development, shaping long-term health, wellbeing, and economic prosperity (Hahn & Truman, 2015). These domains are particularly critical during the transition from adolescence to adulthood, a developmental period in which pathways are often set for future life chances. The category of young people not in education, employment or training (NEET) is used internationally as a marker of youth disengagement and lost productivity. Globally, NEET incidence has been increasing yearly with 20.4% of young people aged 15-24 years NEET in 2024 (International Labour Organization, 2024); with a corresponding estimate of 13.2% among 16-24 year olds in the UK (Office for National Statistics (ONS), 2024).

The long-term consequences of youth NEET are well established, including poorer health outcomes, lower lifetime earnings, and broader economic cost to society (Bynner & Parsons, 2002; Feng et al., 2018; Rahmani & Groot, 2023). A wide range of risk factors including individual, familial, societal, and structural factors such as poverty have been shown to increase the likelihood of becoming NEET (Thompson, 2011). Mental health issues and psychosocial problems in adolescence are also consistently associated with increased risk of NEET (Gariépy et al., 2022; Rahmani & Groot, 2023; Rodwell et al., 2018). Structural barriers such as social exclusion and labour market demands further constrain access to opportunities, disproportionately affecting the most disadvantaged, particularly those with pre-existing mental health difficulties (Boardman, 2011; Thompson, 2011). These risk factors often interact and accumulate over the life course, suggesting that meaningful policy responses on NEET should recognise both individual and structural determinants.

Emerging evidence suggests that individual risk factors for NEET may start in early childhood. For instance, lack of school readiness at age 4-5 in England has been linked to later NEET, largely but not solely through academic attainment (Warburton et al., 2024). School readiness also incorporates social and emotional development, key labour market skills (Deming, 2017), which are increasingly recognised as core skills for labour market success (Likhar et al., 2022), independent of cognitive ability. Thus, disentangling the contributions of these developmental issues could help inform the type and timing of interventions throughout children’s development, beyond academic attainment, which may impact NEET status.

Previous studies have shown that patterns of cognitive and socioemotional development across childhood may impact health and educational outcomes in adolescence (Morris et al., 2018; Pearce et al., 2016), with the timing of emergence of problems to be a crucial factor. For example, our previous research has shown that children with early socioemotional and cognitive problems, where socioemotional problems resolve and cognitive problems improve from age 3-14 years did not experience adverse health outcomes in adolescence (Black et al., 2023), but remained at increased risk of poor educational attainment at age 16 (Black et al., 2025). This distinction in the impact of early and resolved behavioural problems (and reducing cognitive problems), which is positive for health but negative for education, provides an opportunity to understand whether good social and emotional skills (despite some cognitive problems) are also protective for not becoming NEET. Furthermore, quantifying associations between trajectories of socioemotional and cognitive development, which emerge early, later or are persistent, and NEET status provides an opportunity to better understand the balance between the impact and timing of problems in socioemotional and cognitive development on NEET status. Therefore, this study aimed to assess the impact of concurrent cognitive and socioemotional development trajectories in childhood on NEET status at age 17 years in a UK cohort.

## Method

### Study settings and participants

We used longitudinal data from the UK Millennium Cohort Study (MCS). MCS is a large-scale, population-based cohort study tracking over 18 000 children born in the UK between September 2000 and January 2002, with follow up at ages 3, 5, 7, 11, 14 and 17 years, corresponding to waves 1 to 7. The numbers of responding families at the different waves were 18,552 (wave 1), 15,590 (wave 2), 15,246 (wave 3), 13,857 (wave 4), 13,287 (wave 5), 11,726 (wave 6) and 10,625 (wave 7). MCS collects information on various topics at each wave, including education, employment health, family socio-economic circumstances and structure, cohort members’ behavioural and cognitive development. Information was usually provided by the primary caregiver, mostly the cohort members’ mother. However, parental involvement was minimal in wave 7 when the child was 17 years. Detailed information on the survey design, sampling and the scope of the MCS is detailed elsewhere (Connelly & Platt, 2014). Ethical approval for different waves of the study was received through the National Health Service (NHS) Research Ethics Committee (REC) system (Shepherd & Gilbert, 2019), and no additional ethical approval is required for this study.

### Exposures

Our main exposures were trajectories of cognitive and socioemotional development from early childhood to adolescence (age 3 to 14 years) as previously defined in Black et al. (10). Four trajectory groups of child cognitive and socioemotional development experienced by children in the UK millennium cohort were identified using a group-based multi-trajectory modelling approach (Nagin et al., 2018). Socioemotional behaviour was measured using the Strengths and Difficulties Questionnaire (Goodman, 1997), completed by the parent, while cognitive development was measured from the results of standard cognition tests administered individually to cohort members at ages 3, 5, 7, 11 and 14 years (the list and description of measures are summarised in supplementary appendix). The four trajectory groups, based on predicted probabilities, were ‘no problems’ (76.5%); ‘Early and resolving cognitive and socioemotional problems’ (8.6%); ‘late socioemotional problems’ (10.1%); and ‘persistent cognitive and socioemotional problems’ (4.8%) (see Supplementary file - Figure S1).

### Outcomes

The primary outcome was NEET status at age 17. We measure NEET using a binary construct from responses to four questions related to NEET status. Cohort members were asked the following questions in wave 7 of the MCS: (i) “Are you currently going to school or college?”, (ii) “Are you currently doing an Apprenticeship?”, (iii) “Are you currently doing any kind of traineeship, training course or scheme?”, and (iv) “Are you currently doing any kind of paid job? We categorised participants as ‘NEET’ (coded 1), if they responded “no” to all the four questions, and those who responded “yes” in an any of the four questions were classified as ‘Not NEET’ (coded 0).

The secondary outcome was economically inactive NEET. Young people who were NEET may be either unemployed and actively looking for work or economically inactive. Economic inactivity was defined as not being in education, employment, or training, and not actively looking for work in the four weeks prior to the interview (Office for National Statistics (ONS), 2024). This was based on responses to two questions regarding current job search activity and any attempt to find paid job including both employee or being self-employed within the past four weeks(Fitzsimons et al., 2020). Participants who were NEET and reported no active job search during this period were classified as economically inactive NEET, (coded as 1) and otherwise coded as 0.

### Confounders

We adjusted for potential confounders associated with child development in early-life and NEET, guided by a directed acyclic graph (Figure 1). The key confounders considered include child sex, maternal education level (degree or higher, diploma, A-levels, GCSE A–C, GCSE D–G, or no qualifications), maternal ethnicity (white, mixed, Indian, Pakistani and Bangladeshi, Black or Black British, or other ethnic groups), and income quintile at baseline.

**Figure 1:**
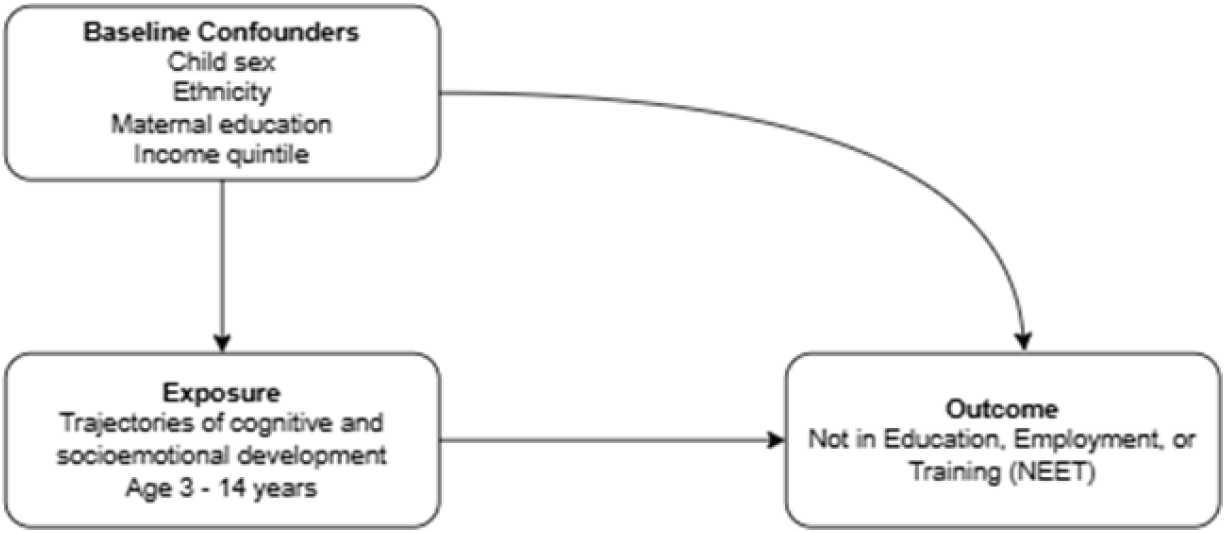
Directed Acyclic Graph (DAG) for the study.

### Statistical analysis

We first described the distribution of sample characteristics including exposures, confounders and outcomes across the trajectories of cognitive and socioemotional behaviour using descriptive statistics. We then estimated Poisson regression models to examine the association between the identified trajectory groups and NEET status at age 17. Two models were constructed: unadjusted model and model adjusted for confounders. We generated risk ratios with robust standard errors comparing each developmental trajectory groups to “no problem” trajectory group. All estimates were reported with 95% confidence intervals and were weighted using longitudinal weights to account for attrition, non-response and sampling design. To assess the potential population-level impact of developmental trajectories, we calculated population attributable fractions (PAF) (Mansournia & Altman, 2018), which estimates the proportion of NEET cases that could be prevented if exposure to cognitive and socioemotional behavioural problems were reduced to same levels as youth in no problems. We further conducted two robustness checks. First, to account for uncertainty in trajectory group membership, we applied Vermunt18 three-step approach (Vermunt, 2010). Second, we addressed potential bias due to missing covariates data by using multiple imputation by chained equation (MICE) (30 imputed data sets), with results pooled using Rubin’s rules. Statistical analyses were performed using Stata (version 18.5).

## Results

There were 10,625 participants in the MCS when the Cohort Members (CM) were aged 17 years. We included a total of 8,368 CMs in the analysis after excluding those with missing data on the exposure trajectories (Supplementary appendix - Figure S2). The overall prevalence of NEET at age 17 was 3.5% (289 cohort members). Among those classified as NEET, 62% (n=180) were economically active, i.e. looking for or available for work during the interview period, while 38% (n=109) were not economically active (Figure 2). Table 1 shows the characteristics of the cohort members by NEET status. Among participants in the “no problem” trajectory group, NEET prevalence was 2.4%. In contrast, NEET status was more prevalent among those with developmental problems: 13% in the persistent cognitive and socioemotional problems trajectory group, 8% in the late socioemotional problems and 4% in the early cognitive and socioemotional problems group. There was a social gradient to NEET status based on household income quintile and maternal educational qualification. The higher the income quintile and education level, the lower the prevalence of NEET.

**Table 1:** Baseline characteristics and outcome by NEET, observed data.

|  | No NEET<br>(n=8079) | NEET<br>(n=289) |
| --- | --- | --- |
| **Trajectories of socioemotional and cognitive development** |  |  |
| No problems | 6465 (97.6) | 160 (2.4) |
| Early and resolving cognitive and socioemotional problems | 645 (95.6) | 30 (4.4) |
| Late socioemotional problems | 684 (92.2) | 58 (7.8) |
| Persistent cognitive and socioemotional problems | 285 (87.4) | 41 (12.6) |
| **Child's sex** |  |  |
| Male | 3717 (96.6) | 131 (3.4) |
| Female | 4087 (96.6) | 143 (3.4) |
| Missing | 275 (94.8) | 15 (5.2) |
| **Maternal Ethnicity** |  |  |
| White | 6414 (96.5) | 236 (3.5) |
| Mixed | 76 (93.8) | 5 (6.2) |
| Indian | 241 (99.2) | 2 (0.8) |
| Pakistani and Bangladeshi | 640 (97.0) | 20 (3.0) |
| Black or Black British | 262 (97.4) | 7 (2.6) |
| Other ethnic group | 156 (98.7) | 2 (1.3) |
| Missing | 290 (94.5) | 17 (5.5) |
| **Maternal education** |  |  |
| Degree plus | 1820 (99.3) | 13 (0.7) |
| Diploma | 773 (97.5) | 20 (2.5) |
| A-levels | 841 (97.2) | 24 (2.8) |
| GCSE A-C | 2408 (96.0) | 100 (4.0) |
| GCSE D-G | 645 (95.0) | 34 (5.0) |
| None | 1304 (94.2) | 81 (5.8) |
| Missing | 288 (94.4) | 17 (5.6) |
| **Household income quintile** |  |  |
| Lowest quintile | 1337 (93.3) | 96 (6.7) |
| Second quintile | 1478 (94.4) | 88 (5.6) |
| Third quintile | 1490 (96.9) | 48 (3.1) |
| Fourth quintile | 1666 (98.5) | 25 (1.5) |
| Highest quintile | 1810 (99.2) | 15 (0.8) |
| Missing | 298 (94.6) | 17 (5.4) |

**Figure 2:**
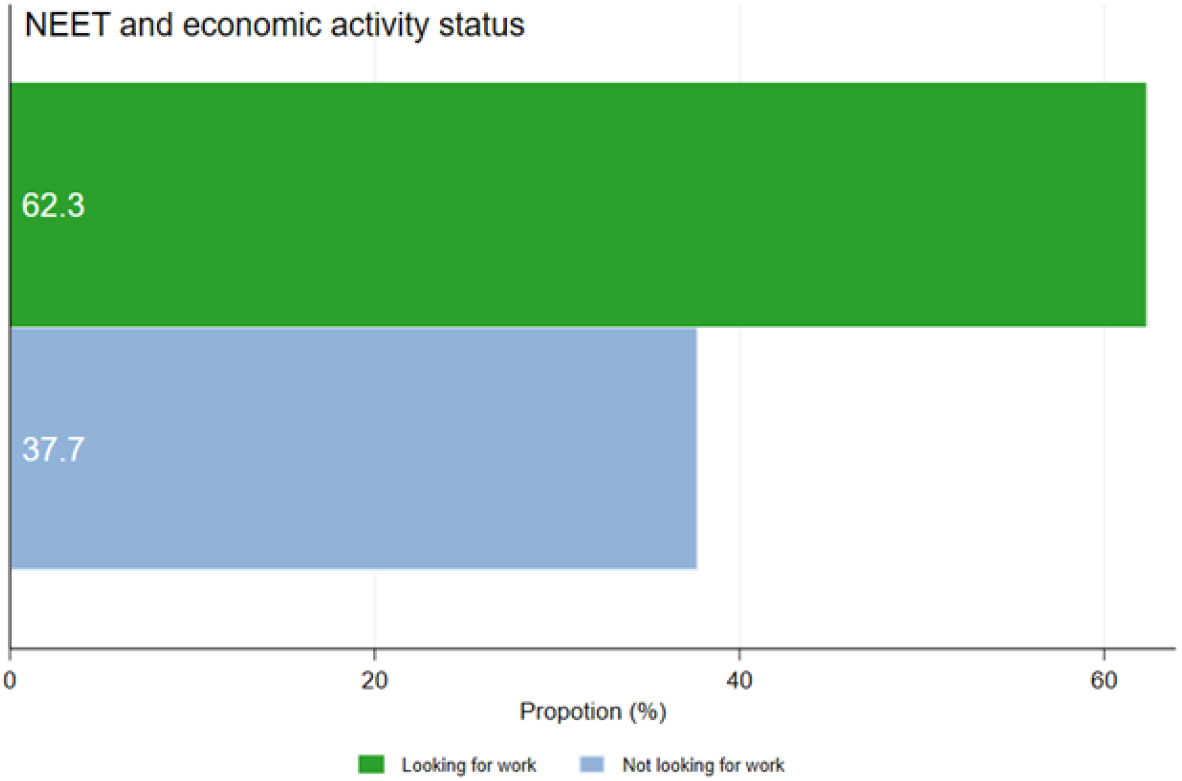
Proportion of NEET in MCS by job search status.

We summarise the estimated association between developmental trajectory groups and NEET status in Table 2 and Figure 3. The unadjusted and adjusted models result both indicate that exposures to early problems were not significantly associated with being NEET at age 17, compared to children in the ‘no developmental problems’ group. In contrast, adolescents with late socioemotional problems were three times more likely to be NEET (adjusted risk ratio [ARR] 3.00, 95% CI 2.08 – 4.32) and those with persistent cognitive and socioemotional problems have three and half times increased risks of being NEET compared to the no problems group (ARR 3.52, 95% CI 2.31 – 5.34). Estimates were slightly attenuated after adjusting for confounders.

**Table 2:**
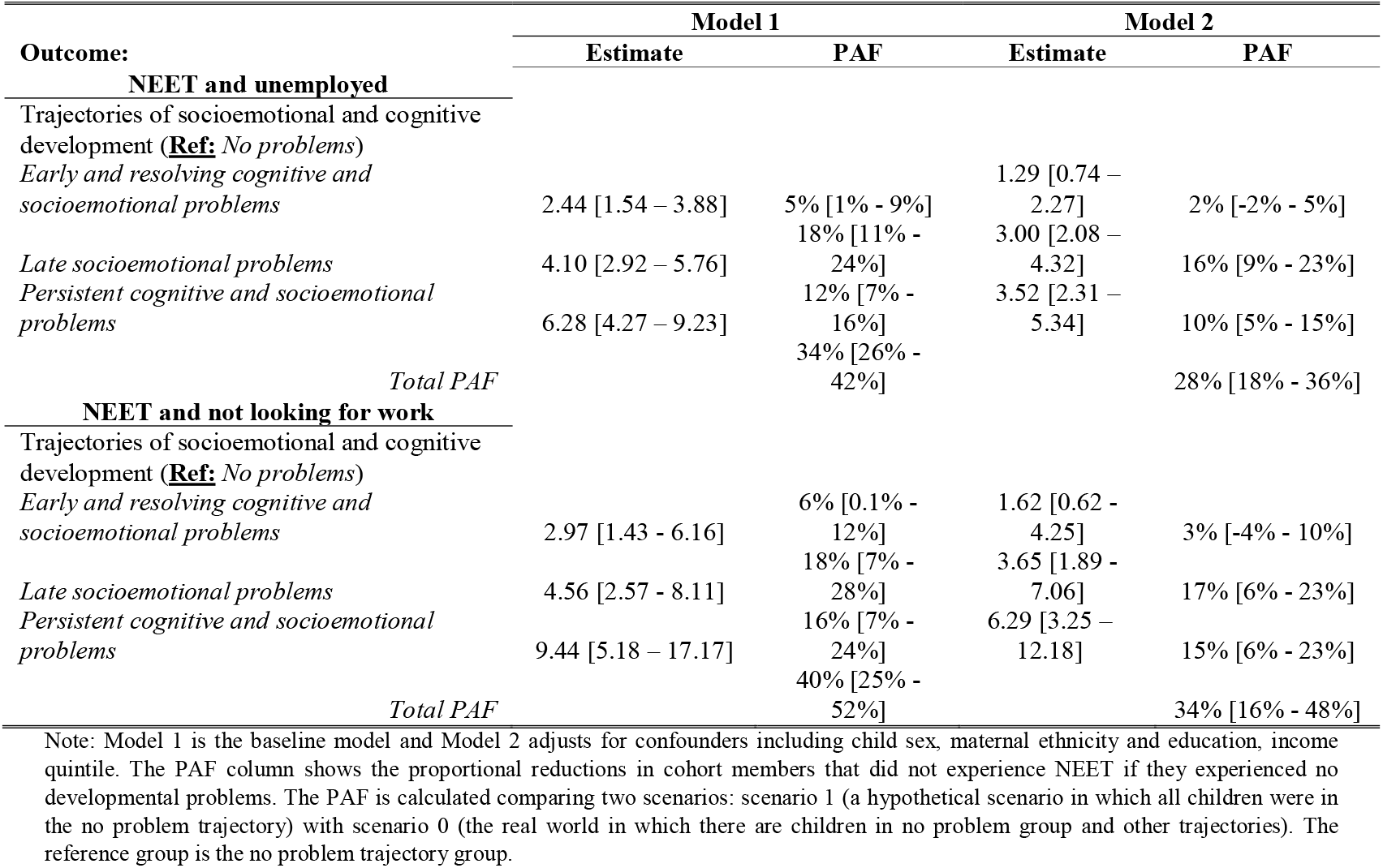
Risk Ratios between multi-development trajectories and adolescents’ NEET in the UK MCS.

**Figure 3:**
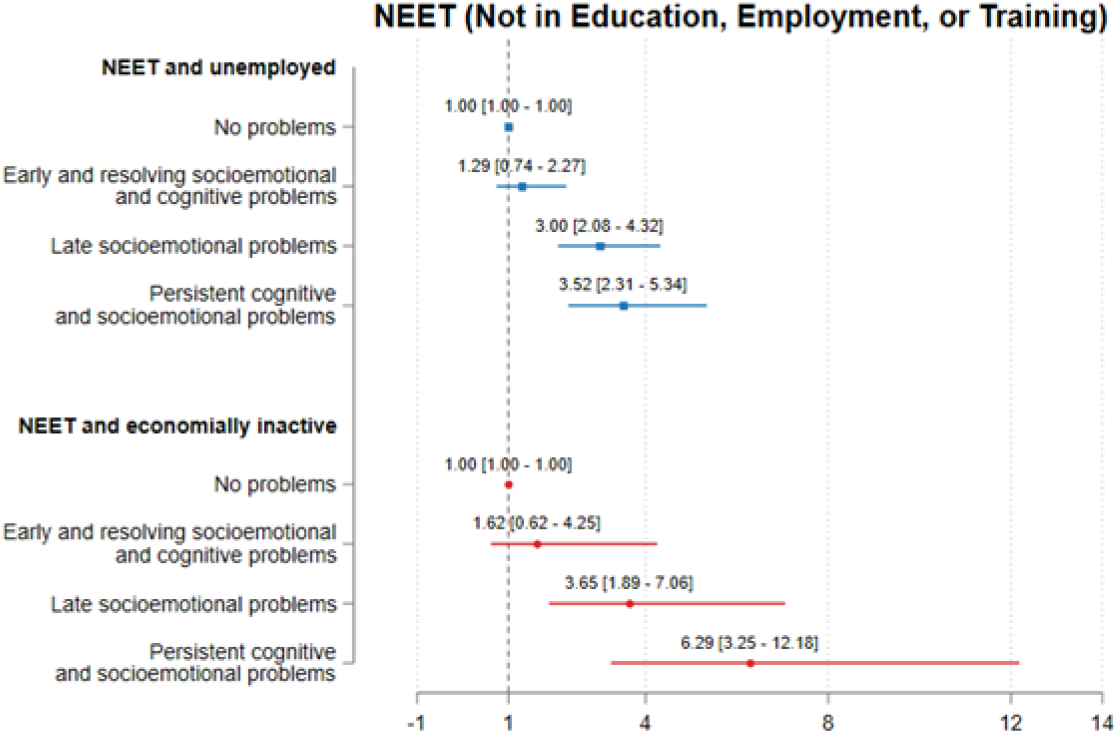
Adjusted risk ratios of predicted developmental trajectories and being NEET at age 17 years in the UK MCS.

Similar patterns were also observed for the secondary outcome (being NEET and economically inactive). Compared to those in the no-problem group, the relative risks were more than three times higher for the late socioemotional problems group (ARR 3.65; 95% CI 1.89 - 7.06) and six times higher for the persistent cognitive and socioemotional behaviour problems group (ARR 6.29; 95% CI, 3.25 – 12.18). Adjustment for confounding factors had minimal impact on the strength of the associations.

The estimated population attributable fractions, which quantify the proportion of NEET cases attributable to each developmental trajectory are presented in Table 2. Assuming a cause– effect relationship, the adjusted PAF shows that approximately 28% (95% CI 18% to 36%) of NEET cases could be attributed to cognitive and socioemotional behaviour problems during childhood. The proportion was higher for adolescents who were NEET and economically inactive, with and adjusted PAF of 34% (95% CI 16% - 48%). The sensitivity analyses, including Vermont three-steps for classification uncertainty and multiple imputation to address missing covariate data show similar results to the main estimation (See Supplementary file – Figures S3 and S4).

## Discussion

We used the Millennium Cohort Study, a large nationally representative cohort sample of UK children, to show the association between joint trajectories of socioemotional and cognitive development throughout childhood and likelihood of not being in education, employment or training in early adolescent years. We found that 3.5% of participants were NEET at age 17, consistent with the national estimate at 3.6% for 16 – 17 years olds in 2017 when CM were 17 years (Office for National Statistics (ONS), 2025). We further showed that approximately 28% of NEET cases were attributable to adverse cognitive and socioemotional development in childhood, if causality is assumed.

Our findings suggest that addressing cognitive and socioemotional behaviour problems during childhood may substantially reduce the risk of becoming NEET in adolescence. Indeed, the timing and persistence of developmental problems were found to be critical. Children with late-onset or persistent problems were at higher risk of NEET, while those with early problems that were on a resolving trajectory did not significantly increase risk compared to children in the ‘no problems’ group.

The distinct role of socioemotional behaviour and its relationship with NEET warrants further investigation. Our results highlight that children who start school with socio-emotional behaviour problems and cognitive problems but are on a resolving trajectory, with behaviour problems resolving by age 7 and cognitive problems reducing, are not associated with NEET status. We know from our other studies that this trajectory group is associated with poor educational attainment at age 16 (Black et al., 2025). So, it appears that resolving socioemotional behaviour problems within the first few years of starting school may be protective of later adverse health and NEET, despite a higher risk of poor educational attainment. This implies that reducing NEET is not limited to exam results but about fostering the social and emotional wellbeing that is necessary for labour market engagement.

Indeed, increasing the wellbeing of working age people is crucial if we are to address the increasing prevalence of economic inactivity (Black et al., 2024). Education, employment and training in young people are pre-cursors for working age economic activity. The current UK government recognises this and one of its missions is to break down barriers to opportunity by expanding high quality education, employment and training to ensure more people are on pathways to good prospects in the next decade (UK Government, 2024). Social mobility policy should also focus on improving routes to employment via apprenticeships and incentivising workplaces to continue with skills development for young people, working across geographical and organisational boundaries and with young people themselves (Public Health England, 2014). Our analysis contributes to understanding the barriers to achieving this goal. Although further research is needed to better understand the mediating pathways, our findings provide strong evidence that NEET status in adolescence is closely linked to earlier cognitive and socioemotional development. Policy efforts need to recognise the importance of developing capabilities and prioritise early intervention strategies that address behavioural and developmental challenges, particularly those emerging or persisting into later childhood (Pemberton, 2008).

Implications beyond social mobility relate to schools and health. Early years investment is crucial to help children’s school readiness, and for those not ready at school entry, schools should be resourced to support development early, ensuring children do not continue to fall behind. The UK government’s focus on school readiness (75% of children should be school ready by 2028) is therefore a welcome and evidence-based policy initiative. Our findings suggest that the emphasis on social and emotional wellbeing in schools is key, with potential benefits for health, employment and welfare. Poor social and emotional wellbeing in childhood is a precursor for poor mental health in later life. If we are to reduce the burden of mental ill health in working age people (Latimer et al., 2025) - a key factor in the rising numbers of young people claiming disability benefit with associated welfare costs spiralling from £36 billion in 2019–20 to £48 billion in 2023–24 (Latimer et al., 2024), then we need to act on social and emotional wellbeing early in life and throughout the child-adolescent life course. This requires a cross-sectoral approach where the health and education sectors work together to support children’s developmental needs (Haug et al., 2023),

To be effective, cross-sector health and education policy needs to be integrated with economic and social policies aimed at mitigating structural disadvantage, in recognition of the contextual factors including increasing child poverty which impact student experience, wellbeing, attendance and attainment. This study adds to a growing evidence linking early developmental trajectories to later educational and economic outcomes (Gariépy et al., 2022; Rahmani & Groot, 2023). Previous research also shows a strong social patterning in developmental pathways with children from socioeconomically disadvantaged backgrounds more likely to follow adverse trajectories (Adjei et al., 2022; Black et al., 2023). Our analyses shows that the association between adverse children development and NEET in adolescence persists after adjusting for key measures of disadvantage, suggesting an independent developmental impact. Therefore, it is imperative to address inadequacies and inequalities in schools funding (Akanni et al., 2024), so that the opportunity to mitigate against disadvantage through the development of capabilities in school is not missed.

The study has some limitations. NEET status was measured at a single time point, which may not capture the dynamic nature of youth transition. Although this is a common limitation (Elsenburg et al., 2025), NEET status at age 17 is of particular relevance given the potential long-term implications for health and employment (Guido, 2015). Also, our analysis did not capture the causality of the relationships between developmental trajectory and NEET. Hence, more causally informed analyses are needed to better understand these relationships. Finally, attrition and missingness are major concerns in longitudinal studies as they pose threats to data quality and could result in estimate bias. Nonetheless, we conducted sensitivity analyses using multiple imputations to address missing data. Our findings are further strengthened using a large contemporary UK birth cohort, enhancing the generalisability and policy relevance of the result. Currently, the England Raising Participation Age legislation, passed in 2013, mandates the participation of young people aged 16 – 17 years in education, training, or employment, recognising the importance of early engagement in these activities.

## Conclusion

Using a representative birth cohort of UK children, we show that adverse cognitive and socioemotional development which persist or emerges late in childhood was strongly associated with increased likelihood of being NEET in adolescence. Coordinated cross-sectoral actions, including health, education and economic systems are needed to address developmental problems. Interventions that target cognitive and socioemotional skills in childhood, particularly in the early years of primary school, may be an effective strategy for reducing the prevalence of NEET and promoting long-term social and economic inclusion.

## Data Availability

All data produced are available online on the UK Data Service Archive

https://datacatalogue.ukdataservice.ac.uk/series/series/2000031?id=2000031#abstract

## Notes

**Financial support** This work was funded by the National Institute for Health and Care Research (NIHR) School for Public Health Research (SPHR), Grant Reference Number PD-SPH-2015. DT-R is also supported by the NIHR on a Research Professorship (NIHR 302438). The views expressed in this publication are those of the authors and not necessarily those of the NIHR or SPHR.

### Competing Interest Statement

The authors have declared no competing interest.

### Funding Statement

This work was funded by the National Institute for Health and Care Research (NIHR) School for Public Health Research (SPHR), Grant Reference Number PD-SPH-2015. DT-R is also supported by the NIHR on a Research Professorship (NIHR 302438). The views expressed in this publication are those of the authors and not necessarily those of the NIHR or SPHR.

### Author Declarations

Ethical approval for data collection for the different waves of the Millennium Cohort Study was approved by the UK National Health Service Research Ethics Committee. Written consent was obtained from all participating parents at each survey; MCS1: South West MREC (MREC/01/6/19); MCS2 and MCS3: London MREC (MREC/03/2/022, 05/MRE02/46); MCS4: Yorkshire MREC (07/MRE03/32); MCS5: Yorkshire and The Humber-Leeds East (11/YH/0203); MCS6: London MREC(13/LO/1786). No additional ethical approval was needed for our secondary data analysis

## References

Adjei, N. K., Schlüter, D. K., Straatmann, V. S., Melis, G., Fleming, K. M., McGovern, R., Howard, L. M., Kaner, E., Wolfe, I., & Taylor-Robinson, D. C. (2022). Quantifying the contribution of poverty and family adversity to adverse child outcomes in the UK: Evidence from the UK Millennium Cohort Study. The Lancet, 400, S16. 10.1016/S0140-6736(22)02226-7

Akanni, L., Taylor-Robinson, D., Hargreaves, D., Creese, H., Barr, B., Ukoumunne, O., Melendez-Torres, G. J., Bennett, D. A., Chua, Y. W., & Esan, O. (2024). OP56 Inequalities in local authority spending on schools in England. A longitudinal ecological analysis 2014–2022. BMJ Publishing Group Ltd. https://jech.bmj.com/content/78/Suppl_1/A22.2.abstract

Black, M., Adjei, N. K., Strong, M., Barnes, A., Jordan, H., & Taylor-Robinson, D. (2023). Trajectories of Child Cognitive and Socioemotional Development and Associations with Adolescent Health in the UK Millennium Cohort Study. The Journal of Pediatrics, 263. 10.1016/j.jpeds.2023.113611

Black, M., Akanni, L., Adjei, N. K., Melendez-Torres, G. J., Hargreaves, D., & Taylor-Robinson, D. (2025). Impact of child socioemotional and cognitive development on exam results in adolescence: Findings from the UK Millennium Cohort Study. Archives of Disease in Childhood, archdischild-2024-327963. 10.1136/archdischild-2024-327963

Black, M., Crawshaw, P., & Barnes, A. (2024). Work and health: We need to focus on people not institutions. Public Health in Practice, 9, 100561. 10.1016/j.puhip.2024.100561

Boardman, J. (2011). Social exclusion and mental health – how people with mental health problems are disadvantaged: An overview. Mental Health and Social Inclusion, 15(3), 112–121. 10.1108/20428301111165690

Bynner, J., & Parsons, S. (2002). Social Exclusion and the Transition from School to Work: The Case of Young People Not in Education, Employment, or Training (NEET). Journal of Vocational Behavior, 60(2), 289–309. 10.1006/jvbe.2001.1868

Connelly, R., & Platt, L. (2014). Cohort profile: UK Millennium Cohort Study (MCS). International Journal of Epidemiology, 43(6), 1719–1725. 10.1093/ije/dyu001

Deming, D. J. (2017). The Growing Importance of Social Skills in the Labor Market*. The Quarterly Journal of Economics, 132(4), 1593–1640. 10.1093/qje/qjx022

Elsenburg, L. K., Kreshpaj, B., Andersen, S. H., De Vries, T. R., Thielen, K., & Rod, N. H. (2025). Childhood adversity trajectories and not being in education, employment, or training during early adulthood: The Danish life course cohort (DANLIFE). Social Science & Medicine, 371, 117841. 10.1016/j.socscimed.2025.117841

Feng, Z., Ralston, K., Everington, D., & Dibben, C. (2018). P10 Long term health effects of NEET experiences: Evidence from scotland. J Epidemiol Community Health, 72(Suppl 1), A65–A66. 10.1136/jech-2018-SSMabstracts.136

Fitzsimons, E., Haselden, L., Smith, K., Gilbert, E., Calderwood, L., Agalioti-Sgompou, V., Veeravalli, S., Silverwood, R., & Ploubidis, G. (2020). Millennium cohort study age 17 sweep (MCS7): User guide. London: UCL Centre for Longitudinal Studies, 1, 97.

Gariépy, G., Danna, S. M., Hawke, L., Henderson, J., & Iyer, S. N. (2022). The mental health of young people who are not in education, employment, or training: A systematic review and meta-analysis. Social Psychiatry and Psychiatric Epidemiology, 57(6), 1107–1121. 10.1007/s00127-021-02212-8

Goodman, R. (1997). The Strengths and Difficulties Questionnaire: A Research Note. Journal of Child Psychology and Psychiatry, 38(5), 581–586. 10.1111/j.1469-7610.1997.tb01545.x

Guido, C. (2015). Young people in transitions: Conditions, indicators and policy implications. To NEET or not to NEET? In Youth and the Crisis. Routledge.

Hahn, R. A., & Truman, B. I. (2015). Education Improves Public Health and Promotes Health Equity. Int J Health Serv, 45(4), 657–678. 10.1177/0020731415585986

Haug, E. H., Nylund, I. B., Samuelsen, N. W., Stokke, M., & Sønderskov, M. (2023). Service Innovations Targeting NEETs: A Systematic Review. Nordic Journal of Transitions, Careers and Guidance, 4(1). 10.16993/njtcg.49

International Labour Organization. (2024). Global Employment Trends for Youth 2024. https://www.ilo.org/publications/major-publications/global-employment-trends-youth-2024

Latimer, E., Pflanz, F., & Waters, T. (2024). Health-related benefit claims post-pandemic: UK trends and global context. The IFS. 10.1920/re.ifs.2024.0333

Latimer, E., Ray-Chaudhuri, S., & Waters, T. (2025). The role of changing health in rising health-related benefit claims. The IFS. 10.1920/re.ifs.2025.0012

Likhar, A., Baghel, P., & Patil, M. (2022). Early Childhood Development and Social Determinants. Cureus, 14(9), e29500. 10.7759/cureus.29500

Mansournia, M. A., & Altman, D. G. (2018). Population attributable fraction. BMJ (Clinical Research Ed.), 360, k757. 10.1136/bmj.k757

Morris, T., Dorling, D., & Smith, G. D. (2018). How well can we predict educational outcomes? Examining the roles of cognitive ability and social position in educational attainment. In Exploring Social Inequality in the 21st Century. Routledge.

Nagin, D. S., Jones, B. L., Passos, V. L., & Tremblay, R. E. (2018). Group-based multi-trajectory modeling. Statistical Methods in Medical Research, 27(7), 2015–2023. 10.1177/0962280216673085

Office for National Statistics (ONS). (2024). Economic activity status variable: Census 2021—Office for National Statistics. https://www.ons.gov.uk/census/census2021dictionary/variablesbytopic/labourmarketvariablescensus2021/economicactivitystatus

Office for National Statistics (ONS). (2025). Young people not in education, employment or training (NEET), UK [Dataset]. https://www.ons.gov.uk/employmentandlabourmarket/peoplenotinwork/unemployment/bulletins/youngpeoplenotineducationemploymentortrainingneet/february2025

Pearce, A., Sawyer, A. C. P., Chittleborough, C. R., Mittinty, M. N., Law, C., & Lynch, J. W. (2016). Do early life cognitive ability and self-regulation skills explain socioeconomic inequalities in academic achievement? An effect decomposition analysis in UK and Australian cohorts. Social Science & Medicine, 165, 108–118. 10.1016/j.socscimed.2016.07.016

Pemberton, S. (2008). Tackling the NEET Generation and the Ability of Policy to Generate a ‘NEET’ Solution—Evidence from the UK. Environment and Planning C: Government and Policy, 26(1), 243–259. 10.1068/c0654

Public Health England. (2014). Evidence review 3: Reducing the number of young people not in employment, education or training (NEET) (Local Action on Health Inequalities: Evidence Papers). https://www.gov.uk/government/publications/local-action-on-health-inequalities-evidence-papers

Rahmani, H., & Groot, W. (2023). Risk Factors of Being a Youth Not in Education, Employment or Training (NEET): A Scoping Review. International Journal of Educational Research, 120, 102198. 10.1016/j.ijer.2023.102198

Rodwell, L., Romaniuk, H., Nilsen, W., Carlin, J. B., Lee, K. J., & Patton, G. C. (2018). Adolescent mental health and behavioural predictors of being NEET: A prospective study of young adults not in employment, education, or training. Psychological Medicine, 48(5), 861–871. 10.1017/S0033291717002434

Shepherd, P., & Gilbert, E. (2019). Millennium Cohort Study: Ethical Review and Consent. https://cls.ucl.ac.uk/wp-content/uploads/2017/07/MCS-Ethical-Approval-and-Consent-2019.pdf#page=5.31

Thompson, R. (2011). Individualisation and social exclusion: The case of young people not in education, employment or training. Oxford Review of Education, 37(6), 785–802. https://www.jstor.org/stable/23120566

UK Government. (2024). Break Down Barriers to Opportunity. https://www.gov.uk/missions/opportunity

Vermunt, J. K. (2010). Latent Class Modeling with Covariates: Two Improved Three-Step Approaches. Political Analysis, 18(4), 450–469. https://econpapers.repec.org/article/cuppolals/v_3a18_3ay_3a2010_3ai_3a04_3ap_3a450-469_5f01.htm

Warburton, M., Wood, M. L., Sohal, K., Wright, J., Mon-Williams, M., & Atkinson, A. L. (2024). Risk of not being in employment, education or training (NEET) in late adolescence is signalled by school readiness measures at 4–5 years. BMC Public Health, 24(1), 1375. 10.1186/s12889-024-18851-w

