## Supplementary material for "Childhood socioemotional and cognitive development and Adolescents NEET (not in education, employment or training): findings from the UK Millennium Cohort Study": Supplentary file

### **Supplementary file**

**Cognitive assessments used at each MCS wave, ages 3-14 years:**

- **MCS2 (Age 3) and MCS3 (Age 5),** BAS Naming Vocabulary: measures expressive verbal ability
- **MCS4 (Age 7),** BAS Word Reading: measures reading ability
- **MCS5 (Age 11),** BAS Verbal Similarities: measures verbal reasoning and verbal knowledge
- **MCS6 (Age 14),** Word Activity Test (subset of the vocab assessment in the 1970 British cohort study survey): measures verbal vocabulary

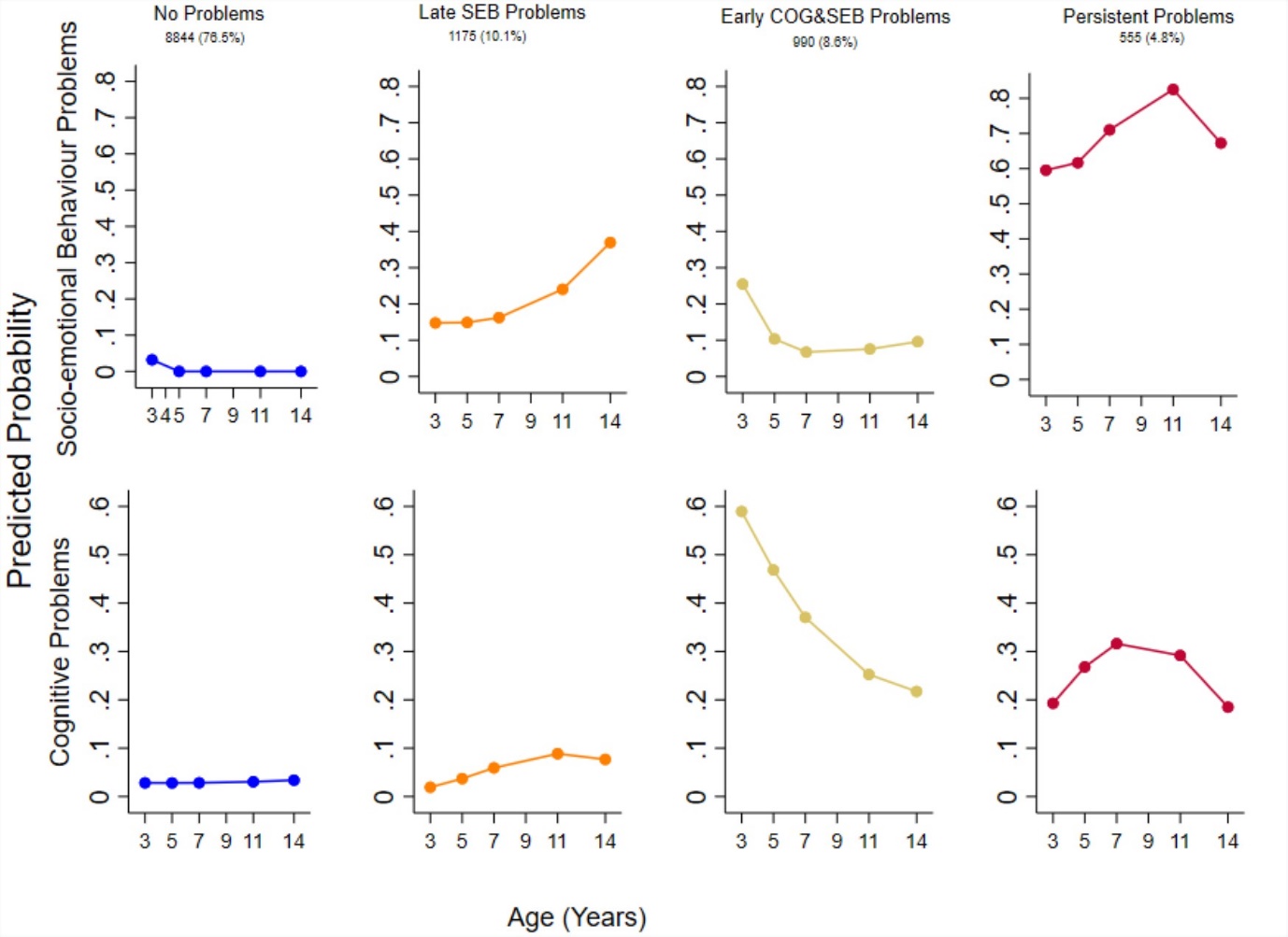

Figure S 1: Developmental trajectory groups by age

Note: the figure depicts the Predicted probability of socioemotional behaviour problems and cognitive problems by age and trajectory group in the Millennium Cohort Study. COG, cognitive development; SEB, socioemotional behavioural development. (source: https://doi.org/10.1016/j.jpeds.2023.113611).

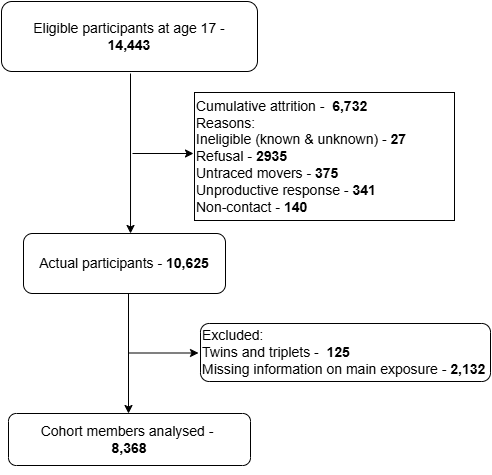

Figure S 2: Study flow diagram showing inclusion and exclusion of cohort participants.

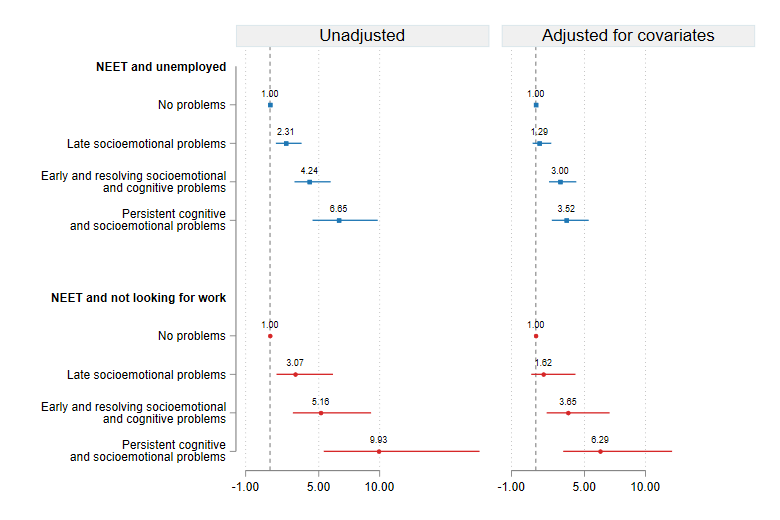

Figure S 3: Trajectory class uncertainties

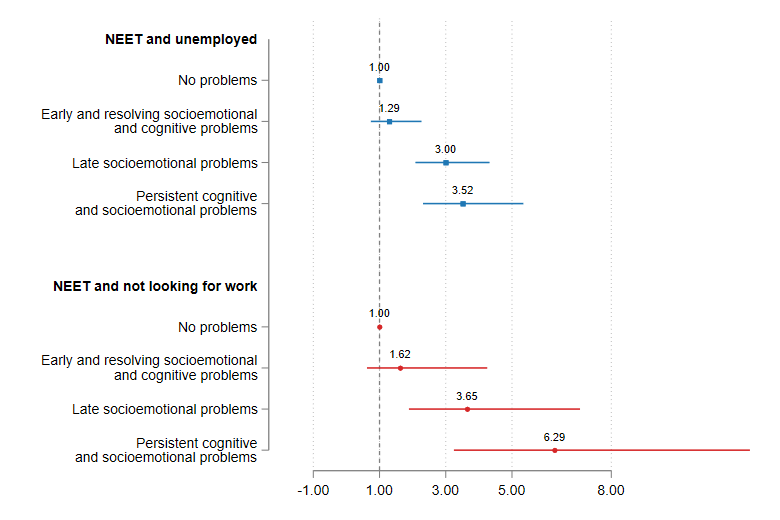

Figure S 4: Multiple imputation results
